# The association of genetically proxied sildenafil with fertility, sexual activity, and wellbeing: a Mendelian randomisation study

**DOI:** 10.1101/2023.03.27.23287822

**Authors:** Benjamin Woolf, Skanda Rajasundaram, Héléne T. Cronjé, James Yarmolinsky, Stephen Burgess, Dipender Gill

## Abstract

**Objective:** To investigate the association of genetically proxied Phosphodiesterase 5 (PDE5) inhibition with fertility, sexual activity, and subjective wellbeing in men.

**Design:** Two-sample *cis*-Mendelian randomisation.

**Setting:** Genetic association data obtained from the International Consortium for Blood Pressure (ICBP) and UK Biobank (UKB).

**Participants:** European ancestry individuals from the ICBP (*N* = 757,601) and the UKB (*N* ≈ 450,000). Genetic association data were leveraged from the ICBP for the exposure and from the UKB for the outcomes.

**Intervention:** Genetically proxied PDE5 inhibition, scaled to the effect of 100mg daily sildenafil on diastolic blood pressure.

**Main outcome measures:** Number of children, age of first having sex, number of sexual partners, odds of being a virgin and self-reported wellbeing, all measured in the male sub-sample of the UKB.

**Secondary outcomes:** To explore the specificity of our results, we replicate our analysis in the female sub-sample of the UKB. We additionally explored possible confounders/mediators of our instruments using PhenoScanner, and adjust for them using Two-step *cis*-MR.

**Results:** Genetically proxied sildenafil was associated with fathering 0.21 (95% CI: 0.08– 0.35) more children (FDR corrected p = 0.01). This association was neither attenuated when adjusting for traits associated with our instruments nor was it replicated in women. We did not find robust evidence for an effect of sildenafil on the age of first having sex, number of sexual partners, odds of being a virgin, or self-reported wellbeing.

**Conclusions:** This study provides genetic support for PDE5 inhibitors increasing the number of children that men have.

**Key Messages:**

- Sildenafil is a PDE5 inhibitor that is commonly used in the treatment of erectile dysfunction and pulmonary hypertension.
- Drug-target Mendelian randomisation is a quasi-experimental method that uses genetic variants to proxy drug-target perturbation. Here, we leverage this approach to investigate long-term therapeutic and adverse effects of sildenafil use, many of which cannot be easily evaluated in a randomised controlled trial.
- We find evidence for a casual association between genetically proxied sildenafil use and number of children fathered. Genetically proxied sildenafil use was not associated with age at first having sex, number of sexual partners, odds of being a virgin, or subjective wellbeing.

## Introduction

Sildenafil (brand names Viagra, Aronix, Liberize, Nipatra, Revatio, Grandipam) is a phosphodiesterase 5 (PDE5) inhibitor commonly used in the treatment of erectile dysfunction (1). PDE5 is an enzyme that promotes the breakdown of the second messenger, cyclic guanosine monophosphate (cGMP), in vascular smooth muscle cells. By inhibiting PDE5, sildenafil increases cGMP activity which induces vascular smooth muscle relaxation and vasodilation. In the setting of erectile dysfunction, this increases blood flow to penile erectile tissue and prolongs erections (2). In the setting of pulmonary hypertension, this induces dilatation of the pulmonary vasculature and improves ventilation/perfusion matching (3). Randomized trials have been effective in identifying various short term adverse effects of PDE5 inhibitors, including indigestion, headaches, flushing, and impaired vision (4,5).

However, while randomised clinical trials (RCTs) provide critical data on drug efficacy, safety, and adverse effects, their limited duration does not allow for the investigation of longer-term secondary health outcomes. For sildenafil, these could include effects on sexual and mental wellbeing, sexual health, and fertility. Now that PDE5 inhibitors are available ‘over the counter’ in countries like the United Kingdom, there may exist repurposing potential for tackling a number of highly prevalent health conditions, including mental health conditions (6) and declining fertility (7).

Investigating these effects using traditional observational studies is undermined by issues of residual confounding and reverse causation. For example, because people who take sildenafil are a non-random subgroup of the general population, any factor which influences someone’s likelihood of taking the medication and which also associates with an outcome of interest could bias the result (e.g., confounding by indication). An increasingly popular epidemiological method for strengthening causal inference in observational settings is Mendelian randomisation (MR) (8,9). Mendel’s laws of inheritance state that genetic variants are inherited independently during meiosis and should therefore not systematically relate to environmental confounding. In the MR paradigm, the random allocation of genetic variants predicting a given exposure at conception is analogous to random allocation to this exposure in an RCT (10). Furthermore, genetic variants are fixed at conception, which confers a greater robustness of MR studies to reverse causation than traditional observational studies and also allows MR studies to estimate the lifetime effects of the exposure.

MR has traditionally been used to study the causal effect of modifiable risk factors, e.g., low-density lipoprotein cholesterol, on clinical outcomes (11). However, given that most drug targets are proteins, and genes encode proteins, MR has been paradigmatically extended to study the effects of perturbing specific drug targets (12). In such drug-target MR studies, variants located within the vicinity of the gene encoding the protein drug target of interest, called ‘*cis*’ variants are used as instruments for drug target perturbation in an instrumental variables (IV) analysis (13). Indeed, *cis*-MR can provide quasi-randomised evidence for outcomes which it might otherwise be infeasible or unethical to investigate using an RCT. For example, a recent *cis*-MR study investigated the efficacy and safety of two major antihypertensive drug classes in pregnancy (14).

This study leveraged *cis*-MR to investigate the long-term effects of genetically proxied PDE5 inhibition on the wellbeing and fertility of men. Specifically, we use variants associated with PDE5A gene expression to mimic the effects of pharmacological PDE5 inhibition on sex-specific fertility (number of children), sexual activity outcomes (age of first having sex, number of sexual partners, and the odds of being a virgin), and subjective wellbeing.

## Methods

### Study design

Two-sample *cis-*MR was performed to explore the association of genetically proxied sildenafil usage with the number of children fathered, age of first having sex, number of sexual partners, odds of being a virgin and self-reported wellbeing in men (Figure 1, Supplementary Figure 1). The *cis* nature of the analysis means that we only considered variants within or near the gene region encoding the drug target as potential instruments. We then used colocalisation and two-step *cis-*MR to explore the robustness of our results to ‘confounding by linkage disequilibrium (LD)’ and/or horizontal pleiotropy (see Table 1 for definitions). Finally, we investigate the sex-specificity of our findings by replicating our analysis in women. A glossary of genetic and Mendelian randomisation specific terms used here can be found in Table 1.

**Figure 1.**
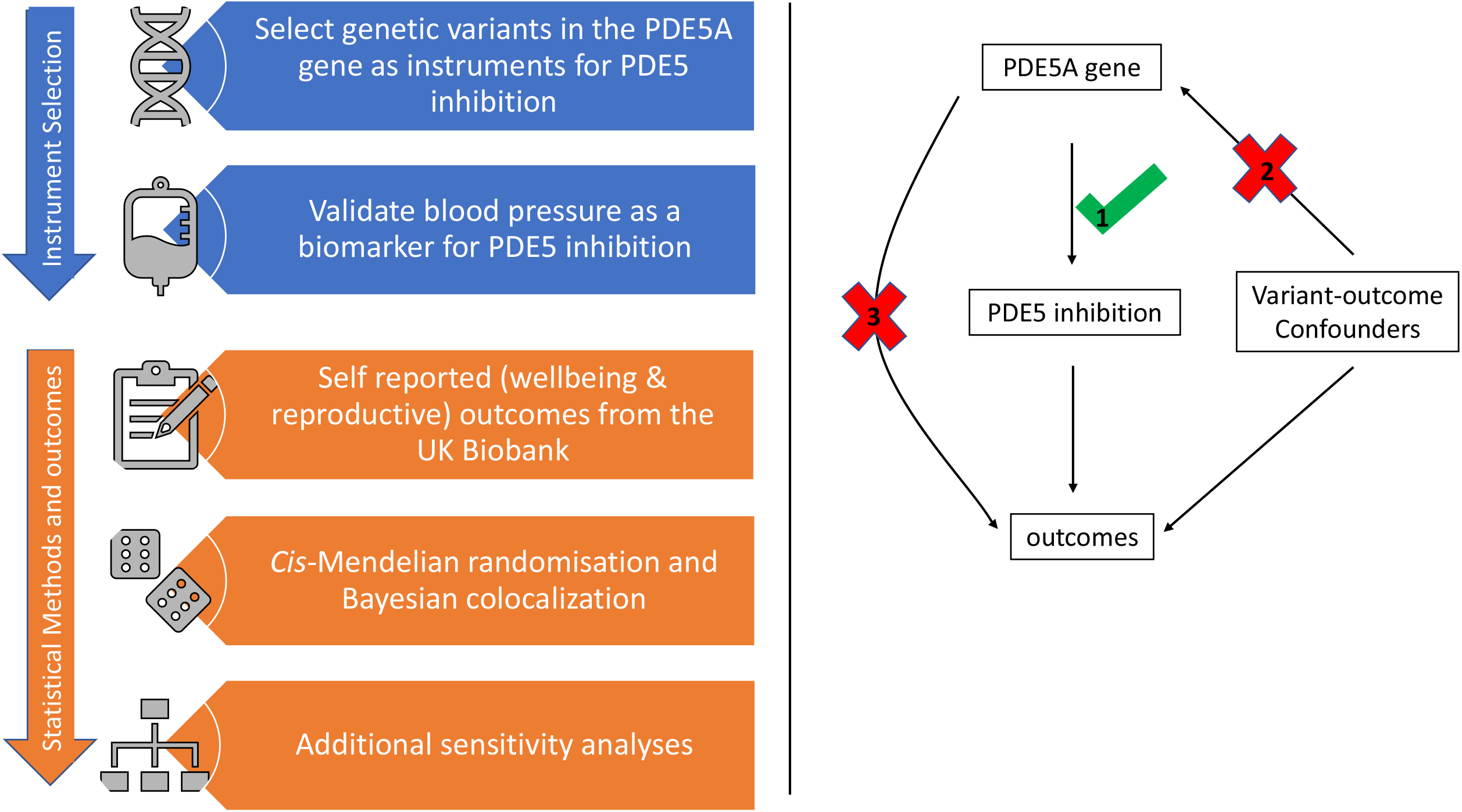
Mendelian randomisation assumes that: (1) the genetic variants in the PDE5A gene associate with PDE5 inhibition (proxied by blood pressure), (2) there are no variant-outcome confounders, and (3) that the variants can cause the outcome only via PDE5 inhibition.

**Table 1:** Glossary of genetics terms

| Term | Definition |
| --- | --- |
| Mendelian randomisation (MR) | A natural experiment which leverages the random allocation of genetic variants (which code for an exposure of interest) at conception to justify an analogy with randomised controlled trials. |
| *cis*-MR | *cis*-MR is distinguished from more traditional ('genome-wide') MR by the use only of variants which are within a gene region of interest. Variants can associate with proxies of drug target perturbation from across the whole genome. However, the mechanism linking these variants is not always clear, so it is typical to restrict drug target MR analyses to variants within or close to the gene region which code for the drug target of interest to ensure an accurate proxy. |
| Linkage disequilibria (LD) | LD occurs when two variants correlate. This occurs because the random shuffling of the parental genotype at birth happens to blocks of variants, rather than individual variants. This means that variants near each other are more likely to be inherited together and hence correlate with each other. LD can be measured either by estimating the correlation between two variants and by looking at the physical number of amino acid bases between them (typically measured with units of kilobases or kb). |
| Confounding by LD | Since variants correlate, MR estimates can be biased if a variant in LD with the instrument affects the outcome of interest through a causal pathway other than via the exposure. |
| Colocalisation | If confounding by LD occurs then the exposure and outcome should not share the same causal variant. Hence, variants which are causal for one trait would not have a similar pattern of associations with both traits. Colocalisation analyses aim to explore this. |
| Genome Wide Association Study (GWAS) | These are large scale population genetics studies which test the association between a phenotype and all measured genetic variants. |
| Pleiotropy | Pleiotropy occurs when genetic variants associate with more than one trait. |
| Horizontal pleiotropy | MR estimates are biased when a variant associates with the outcome of interest independently of the exposure. This type of Pleiotropy is called horizontal pleiotropy. |
| Two-Step *cis*-MR (TSCMR) | A method which uses the principles of mediation analysis to adjust *cis*-MR estimates. Here we use Two-Step *cis*-MR to adjust for potential pleiotropic pathways identified in Phenoscanner. |
| expression Quantitative Trait Loci (eQTL) | These are variants which associate with the expression of a gene. |
| protein Quantitative Trait Loci (pQTL) | These are variants which associate with protein levels. |
| Winner's curse | A bias in genetics studies which occurs if the same data is used to select genetic variants (using a statistical criterion like a p-value) as is used in the analyses. |
| Phenoscanner | A curated database of variant-phenotype associations. |
| Single nucleotide | These occur when there is variation in a single base (i.e. a single letter in the DNA sequence) in more than 0.1% of the |
| polymorphism (SNP) | population. |
| OpenGWAS | A repository of open access GWAS summary statistics |
| BOLT-LMM | A method of analysing GWASs data which uses a linear mixed model to account for confounding by population structure. |

### Data sources

Variant-blood pressure association estimates were extracted from genome-wide association studies (GWASs) by Evangelou *et al*. (2018), on diastolic blood pressure (DBP) and, as a secondary exposure, systolic blood pressure (SBP) (15). This study meta-analysed data from 77 cohorts participating in the International Consortium for Blood Pressure (ICBP) and the UK Biobank (UKB), comprising a total of 757,601 European participants of both sexes. Participating studies measured blood pressure using either manual or automated readings (mmHg) and averaged the two readings where possible. Although the UKB sample, around 60% of the total sample, adjusted for principal components, doing so was optional in the cohorts contributing to the ICBP. All cohorts adjusted for age, age^2^, sex, and BMI, and the UKB data additionally corrected for medication use. Further information on participant and gene level quality control checks can be found in the original publication (15).

Variant-outcome associations were derived from the UKB. The UKB is a large population cohort study of predominantly European ancestry UK residents born between 1934 and 1971 and contains approximately 500,000 participants (16). We conducted male only GWASs for three of the self-reported sexual activity outcomes (age of first having sex: 195,295; number of sexual partners: 203,273; and the odds of being a virgin: 223,568), blood pressure (209,522 DBP and 209,515 for SBP, respectively) and self-reported wellbeing (76,189).

Details of the questionnaires used to obtain self-reported data can be found in the Supplementary Methods. All of our sex-specific UKB GWASs were conducted using BOLT-LMM in the MRC-IEU UKB GWAS pipeline, and adjusted for age, genotyping chip and the first 10 principal components of ancestry (17). A full description of the pipeline methods, including quality control filtering and imputation, can be found in the original citation (17). The pipeline by default excludes participants of the UKB whose genetic sex differs from their reported gender. Variant-outcome information on number of children fathered was extracted from The Elsworth UK Biobank GWAS (OpenGWAS ID: ukb-b-2227, 209,872 males) was extracted from the OpenGWAS repository (18), this GWAS was also conducted using the MRC-IEU UKB GWAS pipeline (19,20). Since around two-thirds of the participants in the Evangelou *et al*. GWAS were from the UKB we expect there to be substantial sample overlap between our exposure and outcome samples.

Further details on data sources used in the secondary analysis, including expression quantitative trait loci (eQTLs) and protein quantitative trait loci (pQTLs), are provided in the Supplementary Methods. The eQTL, pQTL, and Evangelou *et al*. data included both male and female participants.

### Statistical Analysis

#### Instrument selection and validation

To proxy pharmacological inhibition of PDE5, we extracted all *cis* missense single nucleotide polymorphisms (SNPs) identified in PhenoScanner, genome-wide significant pQTLs (SNPs associated with PDE5 protein levels) and genome-wide significant eQTLs (SNPs associated with PDE5A gene expression) from within the PDE5A gene region (GRCh37/hg19 chromosome 4 position 120,415,550 - 120,550,146). To ensure that the variants were not overly correlated with each other, we then ranked these variants in order of the p-values of their associations with DBP as our primary instrumented trait and SBP as a secondary analysis. Variants were then clumped with a LD threshold of r^2^ < 0.35 and a distance threshold of 10,000 kilobases (kB). Since the initial selection of SNPs was not dependent on the statistical significance of their associations with DBP or SBP, this approach is equivalent to ‘three-sample MR’ in terms of its robustness to the Winner’s curse (21).

A two-stage validation process was used for our instrument. First, we performed Hypothesis Prioritisation for multi-trait colocalisation (HyPrColoc) between DBP, SBP, eQTLs and pQTLs to confirm that there was a shared causal variant between them (22). Due to the low coverage of the pQTL GWAS, even with the 200 kB window from the start and end of the gene region used in the HyPrColoc analysis, the HyPrColoc analysis only included 47 SNPs. We therefore additionally implemented ‘LD Check’, as first described by Zheng *et al*., and the details for which are described in more detail in the Supplementary Methods (23). Second, since sildenafil is used routinely in the clinical management of erectile dysfunction and pulmonary hypertension (1,3), we conducted two positive control MR analyses to confirm that our instrument was indeed associated with a reduced liability to these two conditions.

#### Mendelian randomisation

In the MR analysis, the Wald ratio for each genetic variant was estimated by dividing the variant-outcome association by the variant-blood pressure association. These Wald estimates were then meta-analysed with a multiplicative random effects model while using the LD matrix to account for the correlation between variants (24). The Benjamini and Hochberg correction was used to account for multiple testing (25). At a dose of 100mg, sildenafil results in up to an 5.5 mmHg and 8.4 mmHg decrease in DBP and SBP respectively (26,27). To facilitate the interpretation of our results, we scaled our MR estimates to represent the blood pressure-lowering effect of a 100mg daily dose of sildenafil.

### Sensitivity Analyses

#### Colocalisation

One threat to the validity of *cis*-MR analyses is confounding by LD. This occurs when a variant that associates with the exposure is in LD with a variant that associates with the outcome, thereby producing a spurious MR association. To explore the robustness of our results to confounding by LD, we performed Bayesian colocalization using *Coloc*, between DBP and SBP, respectively, and all outcomes for which a significant MR association was identified. *Coloc* presents the evidence for five hypotheses: *H*_*0*_: no causal variant for either trait; *H*_*1*_: causal variant for trait 1 but not trait 2; *H*_*2*_: a causal variant for trait 2 but not trait 1;

*H*_*3*_: distinct causal variants underlying each trait, and *H*_*4*_: a shared causal variant underlying both traits. A high posterior probability (PP) for *H*_*4*_ (PP_H4_ > 0.8) supports the presence of a shared causal variant underlying both traits whereas a high PP_H3_ (PP_H3_ > 0.8) supports the presence of distinct causal variants underlying each trait, and thus indicates confounding by LD in the corresponding MR association. PP_H3_ + PP_H4_ < 0.8 in the presence of a statistically significant MR association implies either a false positive MR finding, or an underpowered Coloc analysis. This will occur when the outcome GWAS is insufficiently powered to detect genome-wide significant associations in the gene region of interest. Since this was the case for many of our outcome GWASs, we used LD Check as a sensitivity analysis for low power.

#### Replication in females

We replicated our primary analyses using GWAS summary data restricted to women to investigate the sex-specificity of our findings. Details on how we ran the female specific outcome GWASs can be found in the Supplementary Methods. Since these GWASs had a similar simple size to the male-only GWASs, they should have similar statistical power.

#### Two-step cis-MR for pleiotropic effects

Horizontal pleiotropy occurs when a genetic variant influences the outcome not solely *via* the exposure, thereby violating the third IV (exclusion-restriction) assumption. To explore potential horizontal pleiotropic effects, we searched Phenoscanner for traits associated with the SNPs included in our instrument at p < 1×10^−5^, as a Bonferroni corrected threshold for the number of traits in Phenoscanner (28). We then used two-step *cis-*MR to adjust our MR estimates for any effect mediated by these traits. Two-step *cis-*MR uses a two-step mediation approach, similar to two-step network MR, to adjust variant-outcome associations for potential pleiotropic pathways or confounding by LD (29,30). We additionally used Two-step *cis-*MR to adjust for body mass index (BMI) to verify that its inclusion as a covariate in the DBP and SBP GWASs had not induced collider bias.

### Patient and Public Involvement

Benjamin Woolf is a pulmonary hypertension patient who has been prescribed PDE5 inhibitors.

## Results

### Instrument validation

Using HyPrColoc, we found a posterior probability of 97% for a shared causal variant between DBP-associated SNPs, SBP-associated SNPs, PDE5 pQTLs and PDE5A eQTLs (Figure 2). This conclusion was supported by the LD Check analysis (Supplementary Table 1). Together, these analyses confirmed the blood pressure as an appropriate biomarker through which genetic instruments of PDE5 inhibition can be identified.

**Figure 2.**
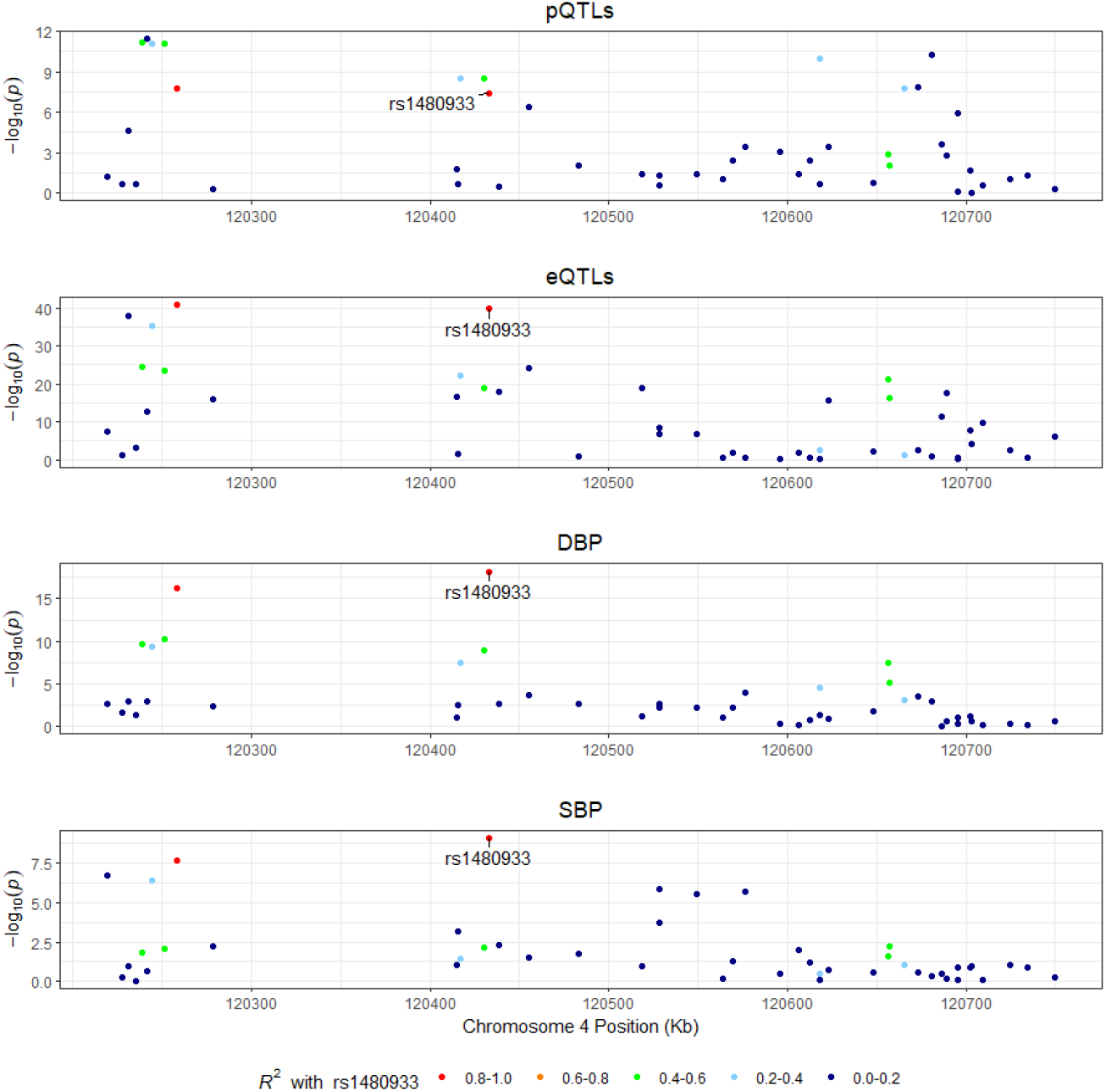
Colocalization analysis for the validation of the genetic instruments, between the two exposures (Systolic and Diastolic Blood Pressure) and gene expression and protein quantitative trait loci (eQTL and pQTL respectively). The putative causal variant in the HyprColoc model is rs1480933.

We instrumented PDE5A-driven DBP and SBP using five (rs80223330, rs12646525, rs17355550, rs10050092, and rs66887589) and four (rs80223330, rs12646525, rs17355550, and rs7672519) variants, respectively. Three variants were represented in both instruments (rs80223330, rs12646525, rs17355550). The average F-statistic was 31.58 for our DBP instrument, and 25.53 for our SBP instrument, indicating low levels of weak instrument bias.

Our positive control analyses yielded an MR association in the expected direction between PDE5 inhibition and erectile dysfunction (p < 0.001 and p = 0.049 for PDE5A-driven DBP and PDE5A-driven SBP respectively) and pulmonary arterial hypertension (p < 0.001 and p = 0.061 and for PDE5A-driven DBP and PDE5A-driven SBP respectively).

### Primary results

#### Number of children fathered

All *cis-*MR estimates of the effect of genetically proxied PDE5A inhibition were scaled to 100mg daily sildenafil use (Figure 3). Our results suggest that PDE5 inhibition is associated with fathering 0.28 more children [95% CI: 0.20–0.36, p_fdr_ < 0.001] when using PDE5A-driven DBP as the instrumented trait. This result was replicated using the PDE5A-driven SBP as the instrumented trait (Table 2). Although the posterior probability of H4 was 3.5 times the posterior probability of H3, H1 was the most likely hypothesis (91%) in our colocalization analysis (Table 2), implying suboptimal power in the colocalisation, which was supported by the low minimum p-value in the GWAS of number of children fathered (Figure 4). Colocalisation was also supported by the LD Check analysis, which found that 68%, 73%, and 55% of the leading 30 SNPs in the DBP, SBP, and number of children fathered GWASs, respectively, were in LD with the SNP most significantly associated with blood pressure.

**Figure 3:**
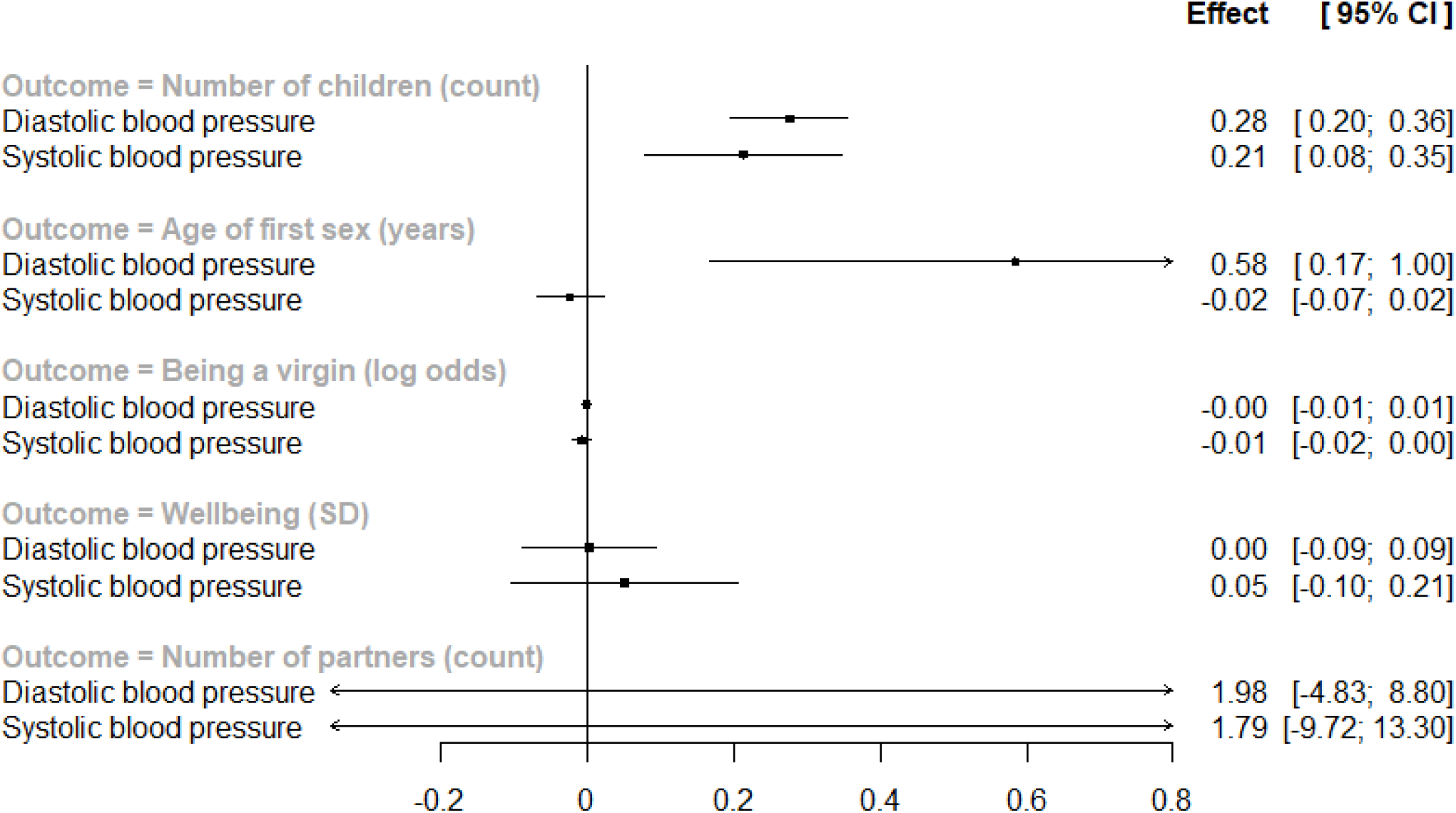
Forest plot of the effect of genetically proxied PDE5 inhibition on the five primary outcomes. Estimates are scaled to the effect of 100mg daily Sildenafil use.

**Table 2:** MR and Colocalization results for the number of children analyses. H0 to H4 represent the posterior probabilities of these hypotheses in a the Bayesian Colocalisation. * This exposure is the primary exposure. MR estimates of the effect of genetically proxied PDE5 inhibition are scaled to the effect of 100mg Sildenafil.

| Analysis |  | Mendelian randomisation |  |  | Colocalisation |  |  |  |  |
| --- | --- | --- | --- | --- | --- | --- | --- | --- | --- |
| PDE5-driven trait instrumented | Outcome | Effect | Lower confidence interval | Upper confidence interval | H0 | H1 | H2 | H3 | H4 |
| Diastolic blood pressure* | Number of children fathered (men) | 0.276 | 0.196 | 0.355 | 0.000 | 0.991 | 0.000 | 0.002 | 0.007 |
| Systolic blood pressure |  | 0.213 | 0.078 | 0.348 | 0.000 | 0.991 | 0.000 | 0.002 | 0.007 |
| Diastolic blood pressure* | Number of live births (women) | -0.136 | -0.277 | 0.005 | 0.000 | 0.996 | 0.000 | 0.002 | 0.002 |
| Systolic blood pressure |  | 0.043 | -0.116 | 0.202 | 0.000 | 0.996 | 0.000 | 0.002 | 0.002 |

**Figure 4:**
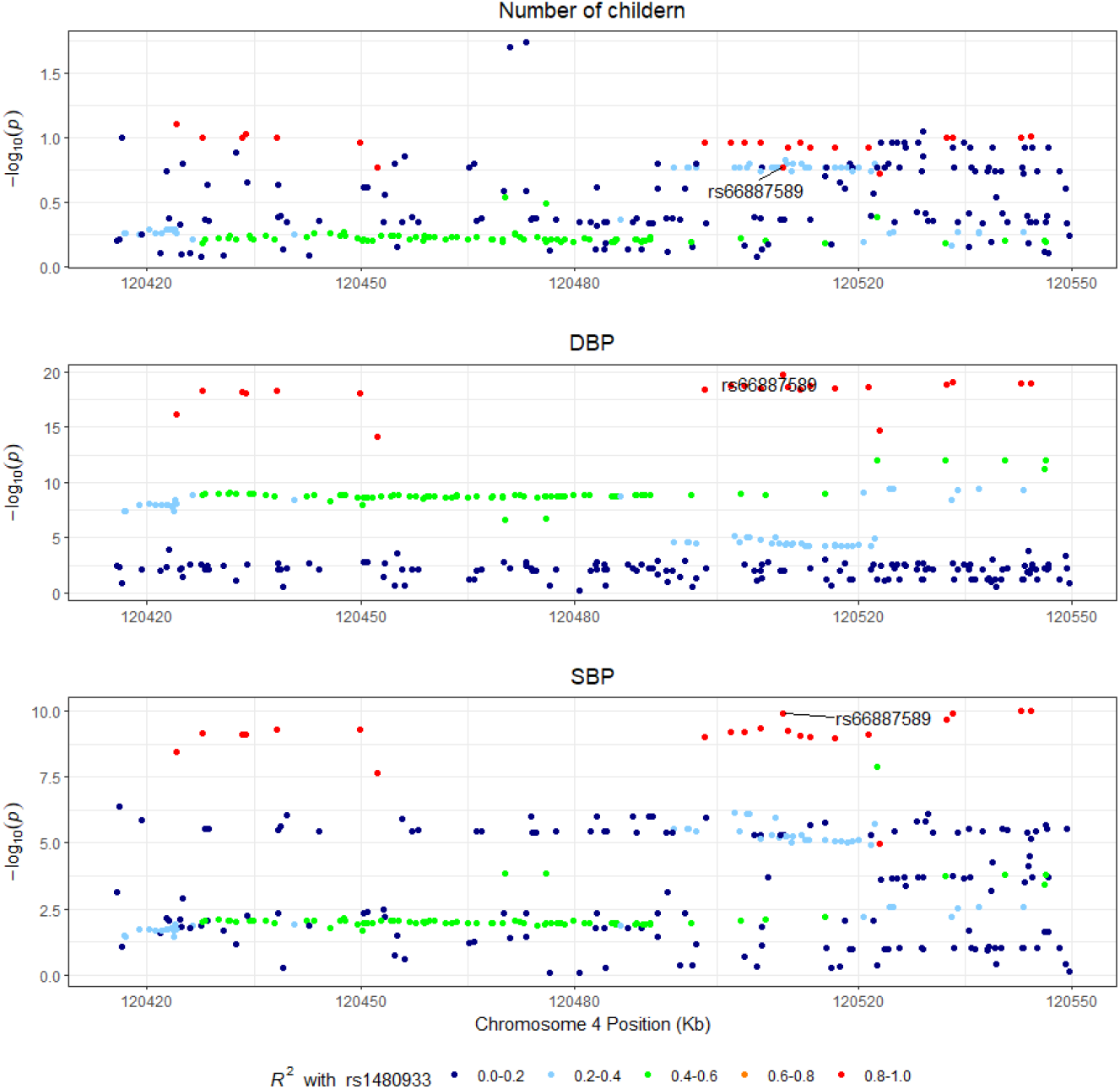
Colocalization plots for the two exposures (Diastolic and Systolic Blood Pressure) with number of children. The putative causal variant in the HyprColoc model is rs66887589.

This conclusion is additionally consistent with the similar pattern in the three locus plots (Figure 4).

#### Age of first having sex

In our primary PDE5A-driven DBP analysis, genetically proxied PDE5 inhibition appeared to influence age (in years) of first having sex (0.584 [95% CI: 168 to 1.000, p_fdr_ = 0.015], Figure 2). However, this result was not replicated using the PDE5A-driven SBP-derived variants (- 0.02 years older, 95% CI: -0.07–0.02, p_fdr_ = 0.933). Further exploration by colocalization analyses and visual inspection of the locus plot indicated the DBP result as a likely false positive finding (PPH1 = 99.6%, PPH3 = PPH4 = 0.2%, Supplementary Figure 3). Furthermore, it failed LD Check since none of the top 30 SNPs associated with the age of first having sex were in LD with the putative causal SNP for blood pressure.

#### Number of sexual partners, odds of being a virgin, and self-reported wellbeing

We found no strong evidence of an association between of genetically proxied PDE5 inhibition, instrumented by PDE5A-driven DBP effects, and any of our other outcomes: the number of sexual partners (1.984 sexual partners [95% CI: -4.829 to 8.797, p_fdr_ = 0.955]), the odds of being a virgin ([odds ratio (OR) = 1.001 [95% CI: 0.992 to 1.005, p_fdr_ = 0.837]), or self-reported wellbeing (standardised mean difference = 0.003 [95% CI: -0.089 to 0.094, p_fdr_ = 0.837]). Similar findings were observed when PDE5 inhibition was instrumented by PDE5A-driven SBP-derived variants (Supplementary Results).

### Sensitivity Analysis

#### Pleiotropic effects

PhenoScanner identified 12 traits which were associated (p < 1 ×10^−5^) with at least one variant included in our instruments (Supplementary Table 2). None of these traits, or BMI, resulted in a meaningful change in effect when adjusting our MR effect estimate for them using two-step *cis*-MR (Supplementary Table 3).

#### Sex-specificity

We did not find evidence of an association between genetically proxied sildenafil and the number of live births in women (β = 0.14, 95% CI: -0.28 to 0.01), although the estimate was in the opposite direction to that in men. Sex-stratified locus plots of DBP and SBP GWASs imply that this result is not due to PDE5A expression only occurring in men (Supplementary Figures 3 and 4). Female only results for the other outcomes are presented in the Supplementary Results, but do not show consistent evidence of an association.

#### Same population assumption

Although all GWASs in our primary analysis were drawn from populations of European adults, we validated the two-sample MR ‘same population’ assumption using the ‘MR Same Population Test’ (31). This failed to find evidence of a difference in association estimates between ICBP and our sex-specific UKB GWASs of blood pressure in the UKB (Supplementary Table 4).

## Discussion

### Key findings and interpretation

In this study, we investigated the association of genetically proxied PDE5 inhibition on male fertility, sexual activity, and wellbeing. Our *cis-*MR analysis found that genetically proxied PDE5 inhibition was associated with fathering more children. Although this MR association was not supported by Bayesian colocalization analyses, it was supported by the LD Check colocalisation analysis and appeared to show a similar pattern on the locus plot, implying that coloc may be underpowered. We did not find evidence of an association with age of first having sex, number of sexual partners, odds of being a virgin or wellbeing.

Our study provides genetic support for a beneficial effect of sildenafil on fertility in men. This is consistent with previous studies which suggest an effect of PDE5 inhibitors on fertility. For example, a systematic review and meta-analysis found that oral PDE5 inhibitors significantly improve sperm motility in men struggling with infertility (32). Similarly, oral PDE5 inhibitors are associated with an increased proportion of morphologically normal sperm in men struggling with infertility, and improved sperm-oocyte binding. PDE5 inhibitors may also indirectly improve fertility by reducing erectile dysfunction (33–36). International Index of Erectile Function (IIEF)-5 scores are significantly reduced in men who are infertile and it has been estimated that more than one third of the men in couples seeking fertility treatment suffer from erectile dysfunction (37,38).

Our sensitivity *cis-*MR analyses did not find an association between genetically proxied PDE5 inhibition and the number of live births for UKB participants, suggesting that our results linking PDE5 inhibition to increased fertility are specific to men. While there exists epidemiological evidence that PDE5 inhibition may have beneficial effects on female fertility and reproductive outcomes (39–44), Cochrane systematic reviews have concluded that such evidence remains inadequate to derive any concrete policy recommendations (45,46). Further studies are warranted in investigating whether PDE5 inhibition exerts an effect on female sexual function that does not translate into live births.

Indeed, a general limitation of population-based studies is the difficulty in appropriately quantifying fertility. The number of children people have is a function not only of the ability to have children but by the desire to have children, amongst a range of other socio-cultural factors. Fertility estimates can be artificially inflated by reproductive assistance, or artificially lowered by contraceptive use, unknown pregnancies, pregnancy termination and/or miscarriages. Although MR is robust to many types of measurement error (47,48), since the prevalence of these factors may not be the same in the UKB as in other studies or populations, our point estimates may not be transferable. Additionally, the absence of relevant and accessible GWAS summary data hindered us from directly investigating the physiological effects of PDE5 inhibition on sexual (dys)function in women using MR.

#### Implications

In this study, we find evidence of an association between genetically proxied sildenafil effect and the number of children fathered. As such, the public health implication of our research is that, at a population level, widespread use of sildenafil and other PDE5 inhibitors could be used to improve fertility outcomes. Since fertility is declining in many countries (49), such an intervention could help reverse this trend.

A second potential implication is that male sexual dysfunction is either under-diagnosed and/or under treated in the UKB and potentially the UK more generally. Assuming that the effect on fertility of PDE5 inhibitors is primarily through its effects on erectile function, our results imply that there may be some residual ED in the study population. This could occur either because people who are diagnosed with ED do not receive treatment, or because some people are not diagnosed with it. Indeed, random samples of general population generally report higher age-specific estimates of erectile dysfunction than the UKB (<3%) (50). The UKB may also have been under treated when compared to a modern cohort. Since sildenafil was only approved for treating erectile dysfunction in 1998, most UKB participants would have already attempted to have had children before access to the drug was widely available.

Literal interpretations of MR estimates can be misleading, especially in instances where the causal estimate is likely to vary across the life course (51,52). Thus, further research is required to estimate how PDE5 inhibitor use may affect the number of children an individual has. Given the wide confidence intervals for our estimates in women, additional studies will also be necessary to further investigate the possibility that PDE5 inhibition in women has a genuine negative effect on the number of children mothered.

### Strength and limitations

To our knowledge, this is the first study to leverage *cis-*MR to investigate the causal effects of PDE5 inhibition. The novelty of our research question is strengthened by our analytical approach that is more robust to the influence of confounding and reverse causation as compared with those derived from traditional observational methods. We demonstrated the validity of our instrument with two positive controls, erectile dysfunction, and pulmonary hypertension and by showing evidence of colocalisation across *PDE5A* gene expression, PDE5 protein expression, DBP and SBP in the *PDE5A* gene region. Furthermore, the presence of strong instruments, and the use of two-step *cis*-MR to adjust for potential pleiotropic pathways provides further assurance that the MR assumptions are valid.

A potential limitation of our study is the apparent failure to replicate the association between genetically proxied sildenafil and number of children fathered in the Bayesian colocalisation analysis. Evidence of colocalization helps ensure that *cis*-MR associations are not arising due to confounding by LD. Despite the absence of supportive evidence from coloc, we believe it is unlikely that our results are attributable to confounding by LD. Firstly, the highest posterior probability was for H1 (only an effect in the exposure GWAS) rather than H3 (confounding by LD), and the posterior probability for H4 (colocalisation) was over three times more likely than H3. The high H1 strongly implies that the outcome GWAS was underpowered. Consistent with this, the smallest p-value in the outcome GWAS is large with respect to the exposure GWAS and locus plots indicate the presence of a similar, albeit deflated, peak around the lead variant in the number of children fathered GWAS as in the blood pressure GWASs. Furthermore, the less conservative LD Check analysis provides supportive evidence for colocalisation.

A second, and important, limitation is generalisability. Since genetic variants are inherited at conception and sildenafil use typically starts post-puberty, the effects estimates derived here may not be completely representative of sildenafil use in practice. Furthermore, our study used European ancestry participants and so we cannot be certain that our results would generalise to other ancestries. Finally, given the availability of only summary level GWAS data, we must assume linearity and cannot explore potential nonlinear effects.

## Summary and conclusions

We find genetic evidence to support an effect of PDE5 inhibition on men having more children. This suggests that a more widespread use of PDE5 inhibitors, like sildenafil, could help alleviate the declining fertility rates in many countries, although further studies are required to prove this definitively.

## Data Availability

The data that was used in this study is publicly available from the MRC-IEU OpenGWAS platform. The R code used in the manuscript, and GWASs created for this project, are available from https://doi.org/10.17605/OSF.IO/MUERZ.

## Other information

### Author contributions

BW designed and implemented the study under the supervision of DG. All authors contributed to the writing of the manuscript.

### Data and data sharing

The data that was used in this study is publicly available from the MRC-IEU OpenGWAS platform. The R code used in the manuscript, and GWASs created for this project, are available from https://doi.org/10.17605/OSF.IO/MUERZ. This work was carried out using the computational facilities of the Advanced Computing Research Centre, University of Bristol - http://www.bris.ac.uk/acrc/. All GWAS summary data used in this research, including that created for the applied examples, will be made publicly available.

This project was conducted using UK Biobank application no. 15825. UK Biobank was established by the Wellcome Trust medical charity, Medical Research Council, Department of Health, Scottish Government and the Northwest Regional Development Agency. It has also had funding from the Welsh Government, British Heart Foundation, Cancer Research UK and Diabetes UK. UK Biobank is supported by the National Health Service (NHS). UK Biobank is open to bona fide researchers anywhere in the world.

### Ethics Approval statement

UKB received ethics approval from the North West Multi-Centre Research Ethics Committee (REC reference 11/NW/0382). All participants provided written informed consent to participate in the study. Data from the UKB are fully anonymised.

