## Supplement for "The association of genetically proxied sildenafil with fertility, sexual activity, and wellbeing: a Mendelian randomisation study"

ICBP SBP or DBP GWAS (757,601 participants, 7,160,619 SNPs)

4 and 5 *cis* SNPs for SBP and DBP respectively after clumping (r^2^ = 0.35 and KB = 10000).

Wellbeing GWAS (76,189 men)

4 missense variants, 16 genome-wide significant eQTLs from eQTLGen (N = 31,684) which include the 3 genome-wide significant pQTLs from Suhre et al. (2017) (N = 1,000)

Age first sex GWAS (195,295 men)

Number of children GWAS (209,872 men)

Number of sexual partners GWAS (285,012 men)

Odds of being a virgin GWAS (264,717 men)

Supplementary Figure 1: Flow chart of SNPs and participants in the primary analysis.

**Supplementary Methods**

Assumptions of two-sample Mendelian randomization, and their assessment

Mendelian randomisation, as a type of instrumental variables (IV) analysis makes three core assumptions: 1) that the instrument (in this context a genetic variant) is strongly associated with the exposure, 2) that the instrument causes the outcome only via the exposure, and 3) that there is no instrument-outcome confounding.

Bias due to violations of the first assumption is inversely proportional to the F statistic for the variant-exposure association. We therefore evaluate this assumption by calculating the F statistic as the square of the variant-exposure association divided by the square of the standard error of this association. The second two assumptions cannot be proven empirically. In a cis-MR setting, the second assumption is plausible since the exposure of interest, PDE5 inhibition, is very proximal to the gene, and there therefore are not many plausible pathways through which a polytopic effect could violate this assumption. We additionally use two-step cis-MR to adjust our MR estimates for traits known to associate with the variants of interest. The third assumption is plausible because of Mendel’s laws of independent and random segregation of genetic variants. A threat to this assumptions validity in a cis-MR setting is confounding by LD (i.e. when a variant in LD with the causal variant independently causes the outcome and therefore introduces confounding). We used colocalization to test for the presence of confounding by LD. The 95% CI in both the two-step cis-MR were estimated using bootstrap standard errors with 100,000 repetitions.

To interpret the point estimate of an MR study, a fourth assumption has to be made. Here we assume monocity, i.e. that if the variant-exposure association is positive, then it is positive for every individual. If this this assumption is violated, then MR will estimate the ‘complier average treatment effect’, i.e. the effect in people whose blood pressure is increased by the exposure, given an increase on average. Since GWASs assume linear models, our MR analysis will additionally assume linearity. If this assumption is violated then MR will still provide a valid estimate of the average causal effect (1).

By using two-sample MR we additionally need to assume that the samples are drawn from a comparable population. To do this we ensure that the exposure and outcome samples are form the same ethnicity and age range. In addition, we apply the MRSamePopTest falsification test (2). This tests for homogeneity in the variant-exposure associations, and therefore tests the exchangeability of effect modifiers assumption required by the potential outcomes framework for an effect estimate to be generalisable form one setting to another. In addition, to the extent that there is no sample overlap in MR studies, weak instrument bias will bias estimates only towards the null, rather than in either direction. We therefore additionally check for sample overlap between the GWASs.

LD Check

LD Check was introduced by Zheng et al as an alternative method to perform colocalization when there are insufficient SNPs to perform a traditional Bayesian ‘coloc’ analysis (3). LD Check is implemented by checking that at least one of the top 30 cis variant is in strong LD (r^2^ = 0.8) with the putative causal variant. These two variants would therefore be in sufficiently strong multi-collinearity that a regression model would not be able to model them as two distinct variables. Here we used the putative causal SNP from the HyPrColoc analysis as the causal variant. More details can be found in the supplement of the original paper.

Female only GWAS

The female only GWASs were conducted using the same methods as the male only GWASs.

To estimate variant-outcome associations in women, we used the female subset of sex-stratified GWASs for SBP (N=247,552). DBP (N=247,558), subjective wellbeing (N=89,815), the age of first having sex (N= 228,579), number of sexual partners: (N=235,926), and the odds of being a virgin: 264,717 females),and systolic blood pressure (247,552 and 247,558 females and diastolic blood pressure respectively). The corresponding estimate to male fertility in women were the number of children they had birthed. This GWAS was performed using UKB data (OpenGWAS ID: ukb-b-1209, N=250,782) (9).

Additional information of UKB phenotyping

Information on wellbeing (UKB ID: 4526) was ascertained through a single question asked in a 2009 follow-up: "In general how happy are you?", and then to choose either: “Extremely happy”, “very happy”, “Moderately happy”, “Moderately unhappy”, “Very unhappy”, “extremely unhappy”, “Do not know”, or “Prefer not to answer”. Information on age of first sex (UKB ID: 2139) was ascertained through a single question asked at recruitment: "What was your age when you first had sexual intercourse? (Sexual intercourse includes vaginal, oral or anal intercourse)". Information on number of sexual partners (UKB ID: 2149) was ascertained through a single question asked at recruitment: "About how many sexual partners have you had in your lifetime?". The odds of being a virgin were then estimated by recoding this question into a binary indicator so that people who had had one or more sexual partner as 1 and leaving those who had had none as coded as zero. Blood pressure was measured using automated readings.

Data sources used in sensitivity analyses

eQTL data was taken from the 2018 eQTLGen Consortium GWAS (OpenGWAS ID: eqtl-a-ENSG00000138735) of whole blood PDE5A expression (4). This study measured gene expression in 31,684 male and female participants of European ancestry. pQTL data was taken from Suhre et al (OpenGWAS ID: prot-c-5256_86_3) blood plasma proteome GWAS (5). This GWAS had 1,000 male and female participants, also of European ancestry.

Associations with erectile dysfunction were estimated as the weighted average meta-analysis of SNP effects form two GWASs. Firstly, the Bovijn et al (2018) GWAS of erectile dysfunction (OpenGWAS ID: ebi-a-GCST006956). This GWAS had 6,175 European cases and 217,630 European controls (6).

Secondly, we used the FinnGen round 8 GWAS of erectile dysfunction (OpenGWAS ID: finn-b-ERECTILE_DYSFUNCTION). This GWAS had 2,038 medical record inferred cases, and 157,478 controls. Finngen is a population cohort study of male and females of Finnish ancestry individuals living in finland (7). Information of pulmonary arterial hypertension was extracted from the FinnGen round 8 GWAS of this trait in the OpenGWAS project (OpenGWAS ID: finn-b-I9_HYPTENSPUL). This GWAS had 213 medical record inferred cases, and 355,864 controls.

Data on impendence of right leg (n = 454,863, OpenGWAS ID: 'ukb-b-7376'), Platelet count (n = 350,474, OpenGWAS ID: ukb-d-30080_irnt), BMI (n = 461,460, OpenGWAS ID: ukb-b-19953), standing height (n = 461,950, OpenGWAS ID: ukb-b-10787), impedance of right arm (n = 454,826, OpenGWAS ID: ukb-b-7859), impedance of left arm (n=454,850, OpenGWAS ID: ukb-b-19379), impedance of whole body (n = 454,840, OpenGWAS ID: ukb-b-19921), impedance of left leg (n = 454,857, OpenGWAS ID: ukb-b-14068), and white blood cell count (OpenGWAS ID: ieu-b-30) were extracted from existing UKB GWASs (8,9).

Myeliod white cell count, Granulocyte count, and Sum basophil neutrophil counts was extracted from the Astle et al (2016) GWAS of that trait (OpenGWAS ID: ebi-a-GCST004626, ebi-a-GCST004614, and ebi-a-GCST004620 respectively) (10). This GWAS had around 170,00 male and female participants, mostly recruited form UK Biobank sub-samples. Coronary artery disease data was taken from the van der Harst et al (2017) GWAS of this trait (OpenGWAS ID: ebi-a-GCST005195). This GWAS had 122,733 cases and 424,528 Controls (male and female, of European ancestry) recruited from the UKB and CARDIoGRAMplusC4D (11).

Software and Preregistration

MR analyses in this paper were run using the TwoSampleMR, TwoStepCisMR, MRPopTest, GGplot, and meta R packages (2,14–18). Some GWAS data was extracted from the MRC-IEU OpenGWAS platform (9). This study was not pre-registered.

**Supplementary Results**

Descriptive data

*Number of participants and SNPs in each sage:*

Figure 1 presents a participants and SNP flow chart. After clumping (r^2^ = 0.35, kb = 10,000) our list of missense variants and eQTLs, our instrument for SBP contained 4 variants (rs7672519, rs80223330, rs12646525, and rs17355550), and the DBP instrument also contained 5 variants (rs17355550, rs12646525, rs80223330, rs10050092, and rs66887589).

*Two-sample MR specific assumptions:* sample overlap and same pop

All GWASs in our primary analysis were drawn from populations of European adults. MRSamePopTest additionally failed to find evidence of a difference in effect between blood pressure measured in ICBP and sex specific GWASs of blood pressure in the UKB (Supplementary Table 1) (28). Finally, there was around 60% sample overlap between the UKB and our exposure GWASs.

*Secondary analysis using Systolic Blood pressure (SBP)*

We replicated our association with Systolic blood pressure (SBP) in our DBP analysis. Our results again suggest that PDE5 inhibition is associated with fathering 0.21 more children [95% CI: 0.08–0.35, p_fdr_ = 0.01].

We again found no strong evidence of an association between of genetically proxied PDE5 inhibition, instrumented by SBP effects, and any of our other outcomes: age of first having sex (-0.02 years, 95% CI: -0.07–0.02, p_fdr_ = 0.93), the number of sexual partners (1.79 more sexual partners, 95% CI: -9.72–13.3, p_fdr_ = 0.87), the odds of being a virgin ([odds ratio (OR) = 0.99, 95% CI: 0.98–1.00, p_fdr_ = 0.93]), or self-reported wellbeing (standardised mean difference = 0.05, 95% CI: -0.10–0.21, p_fdr_ = 0.50]).

*Results in women*

We did not find consistent evidence of an association in the female-only sub-sample of the UKB of PDE5 inhibition with number of sexual partners (DBP: beta = 0.117, se = 0.070, p_fdr_ = 0.156, SBP: beta = 0.106, se = 0.077, p_fdr_ = 0.425), age of first having sex (DBP: beta = 0.086, se = 0.022, p_fdr_ < 0.001; SBP: beta = 0.056, se = 0.025, p_fdr_ = 0.110), log odds of being a virgin (DBP: beta = -0.0003, se = 0.0006, p_fdr_ = 0.692; SBP: beta = 0.0001, se = 0.0006, p_fdr_ = 1.000), or wellbeing (DBP: beta = 0.062, se = 0.022, p_fdr_ = 0.016; SBP: beta = 0.009, se = 0.013, p_fdr_ = 0.759). Please note that these betas are scaled to the effect of a one unit change in mmHg blood pressure on the respective outcomes.

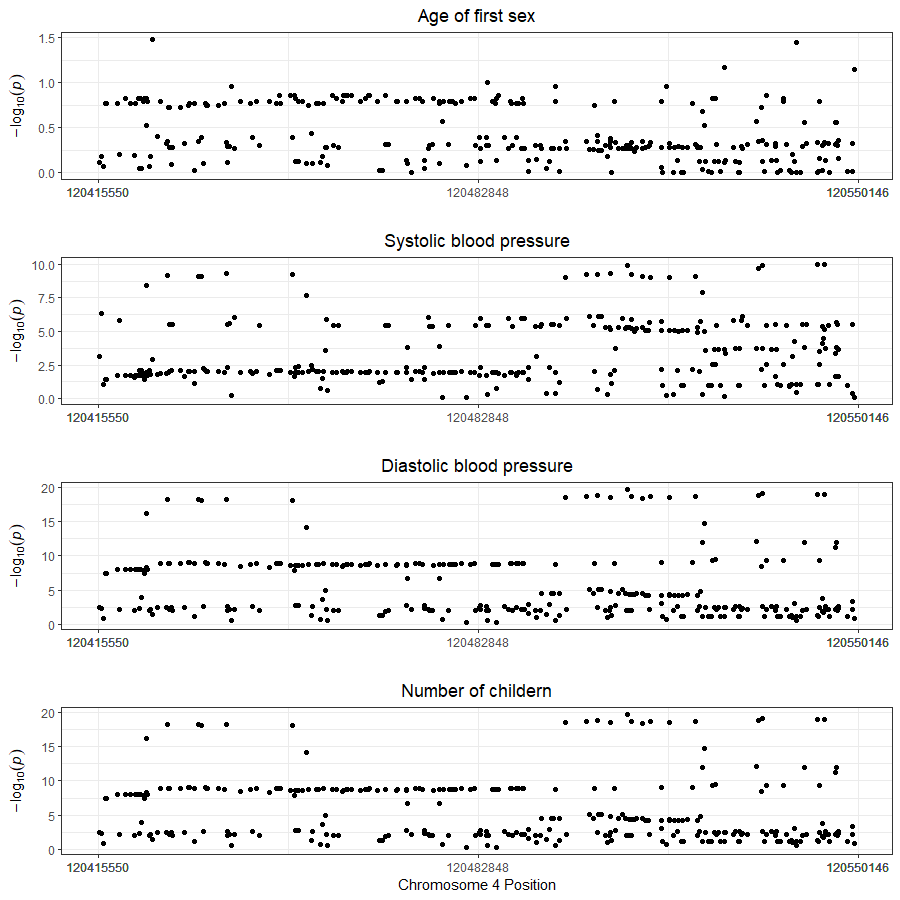

Supplementary Figure 2: Colocalization plots for the two exposures (Systolic and Diastolic Blood Pressure) with age of first sex.

**Supplementary Discussion**

Additional limitations

This study has three key limitations. Firstly, there was large overlap between the exposure and outcome GWASs. Although often described as an ‘assumption’ of two-sample MR, the primary effect of no sample overlap is to ensure that weak instrument bias shifts effect estimates towards the null, thereby reducing the type I error rate. Since, the ICBP GWAS contained UKB participants, there is likely to be a very large amount of sample overlap. However, given the F-statistics of 23 for SBP and 18 for DBP, we would expect the risk of weak instrument bias to be approximately 4% and 6%, respectively. Such bias is unlikely to materially affect the results or conclusions of our study.

A second potential limitation is residual confounding in the GWAS effect estimates. Furthermore, although MR leverages the random allocation of genes during meiosis, the independence of genetic variants with respect to environmental confounding is conditional on parental genotype. At a population level, the inheritance of genetic variants is only approximately random (19). A major source of bias is confounding by ancestry (20). BOLT-LMM, the software used to perform the outcome GWASs, corrects for this bias by adjusting for the entire genetic relationship matrix (a covariance matrix of the number of alleles each participant has in common with every other participant) of all participants. However, the ICBP did not account for confounding by ancestry in all participating GWASs. We would therefore expect our results to be biased towards the null, since we expect greater inflation in the denominator of the MR Wald ratios.

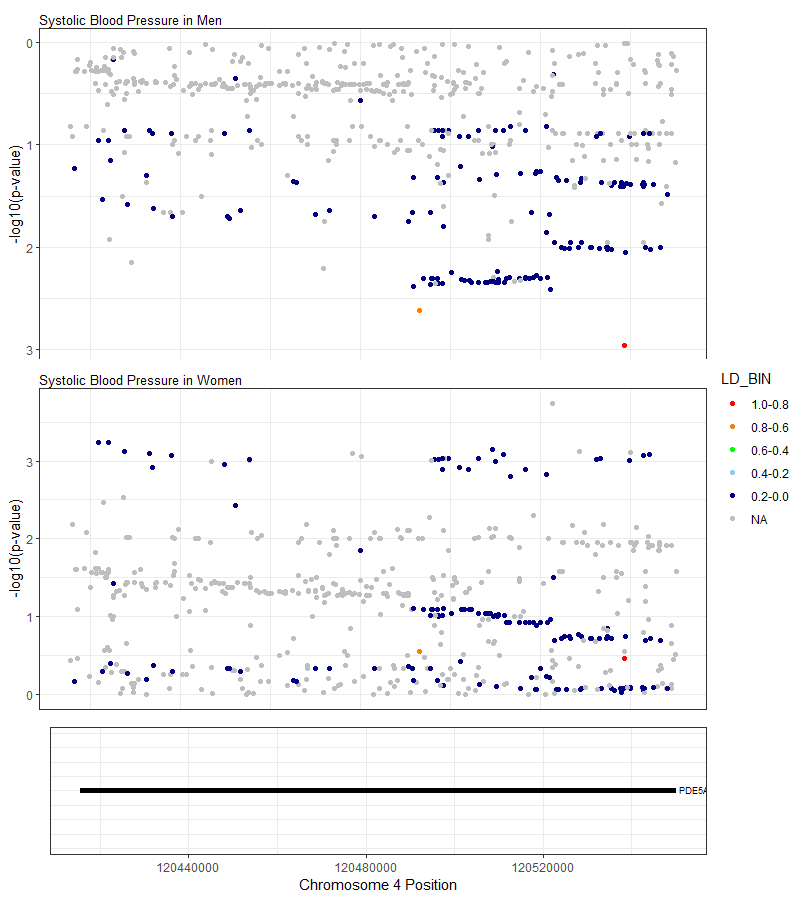

Supplementary Figure 3: Locus plot for Systolic Blood Pressure between male and female only UKB GWASs.

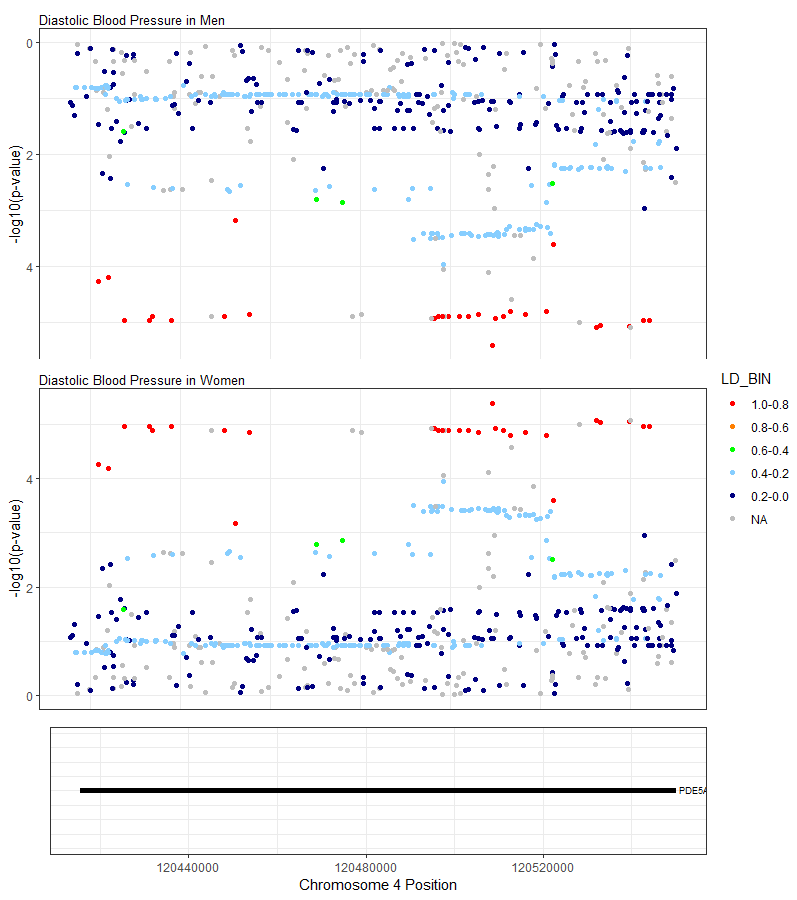

Supplementary Figure 4: Locus plot for Diastolic Blood Pressure between male and female only UKB GWASs.
